# The connection of growth and medication of COVID-19 affected people after 30 days of lock down in India

**DOI:** 10.1101/2020.05.21.20107946

**Authors:** Achintya Bhattacharyya, Debosmita Bhattacharyya, Joy Mukherjee

## Abstract

The COVID-19 pandemic has already consumed few months of indolence all over the world. Almost every part of the world from which the victim of COVID 19 are, have not yet been able to find out a strong way to combat corona virus. Therefore, the main aim is to minimize the spreading of the COVID-19 by detecting most of the affected people during lockdown. Hence, it is necessary to understand what the nature of growth is of spreading of this corona virus with time after almost one month (30 days) of lockdown. In this paper we have developed a very simple mathematical model to describe the growth of spreading of corona virus in human being. This model is based on realistic fact and the statistics we have so far. For controlling the spread of the COVID-19, minimization of the growth with minimum number of days of lockdown is necessary. We have established a relation between the long-term recovery coefficient and the long-term infected coefficient. The growth can be minimized if such condition satisfies. We have also discussed how the different age of the people can be cured by applying different types of medicine. We have presented the data of new cases, recovery and deaths per day to visualize the different coefficient for India and establish our theory. We have also explained how the medicine could be effective to sustain and improve such condition for country having large population like India.

## Introduction

The declaration of pandemic situation due to COVID 19 on 11^th^ March by WHO [1] has made almost all the world to lockdown several countries, either partially or fully to control the spreading the virus. The invisible manifestation of a respiratory infection with symptoms ranging from mild common cold to a severe viral pneumonia leading to acute respiratory distress, is potentially fatal for the people having COPD and other lung deceases. The outbreak of such virus was earlier predicted in China [2]. The present time motivation is to find out as much possible number of such infected cases to slow down the contamination chain of the virus. Hence more and more testing were done during the lockdown period to identify the victims of COVID 19. At the same time, it is also an important that we can identify the maximum number of cases with minimum lockdown days, since the lockdown hampers the economical growth. There are lots of societal impact of lockdown which has already been reported [3]. Hence the question is how to minimize the spreading of CoVID-19? Scientists are trying to develop many statistical and mathematical model [4-5]. Recently a few mathematical models have come across to describe this spread of corona virus to estimate the minimum number of lockdown days to reach base line of the contamination [6-9]. Almost every natural phenomenon follows an exponential law and hence to saturate the graph we must maintain a long lockdown period. At the contemporary time, we see that the number of corona virus infected people are increasing day by day even after 30 days of constant lockdown. Hence to minimize the effect we must need a vaccine to stop the increase of spreading, or we must involve in R&D that which medicine is applicable for faster recovery of patients. No such strong evidence has yet been published so far, that connects the medicine to mathematics.

Here in our paper we propose a simplistic model to visualize the control the covid-19 growth. We have developed the growth equation based on number of active cases, no. of people recovered, and the number of people died. The minimization of the growth rate indicates that we can have the control over COVID 19 epidemic when the coefficient of long-time recovery will dominate over the coefficient of long time inflectibility. We have explained how this can be achieved. For the better understanding we have presented some growth and recovery data to understand the long- and short-term medication effect. In doing this we have collected data from very authentic source worldometer [10]. In our previous work we have shown that a four-degree polynomial fitting for daily new case is a good approximation [11]. Here we have used the same concept to build the growth equation with time. The minimization of the growth equation shows a relation between the coefficients. We have explained this and corelate the effect of medication.

### Mathematical Approach

To construct a growth equation of corona virus affected people, we have taken into consideration of four numbers. The number of corona virus affected people at ant point of time will be denoted by N(t). N_0_ be the number of preexisting active case. The number of daily upcoming case will be denoted by N_1_(t). There are a huge number of people who are recovering daily and let’s say those number is N_2_(t) and finally, N_3_(t) is the number of people dying with time. Now at any instant, we can write an equation which can balance between all the parameters.

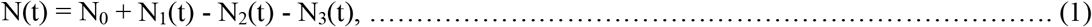

The negative sign of N_2_(t) and N_3_(t) are negative because it reduces the number of COVID-19 patient at any point of time. If any change in the number of COVID-19 patient will be given by the rate of change of N(t), i.e. dN(t)/dt which can be obtained by having derivative of the equation (1).

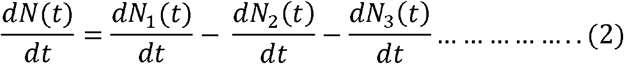

The derivative of N_0_ is put to zero since it is a constant number.

Now currently we introduce a new concept. We know that the growth of natural phenomena increases exponentially with time. Let us assume that the growth of daily reported case, recovered people increases cubic polynomial of time, whereas the number of people who died varies with linear in time (since the mortality rate in India is very less compared to the other countries). Then the N_1_(t), N_2_(t) and N_3_(t) can be written as follows. N_1_(t) = N_10_ + a_1_t +b_1_t^2^ + c_1_t^3^, N_2_(t) = N_20_ + a_2_t +b_2_t^2^ + c_2_t^3,^ N_3_(t) = N_30_ + a_3_t where a_i_, b_i_, c_i_ are the co-efficient of different polynomial term which has different physical significance and ‘t’ is the number of days spent after lockdown announcement. These assumptions are based on completely realistic point of view. The coefficient plays important roles in controlling the spread of corona virus. If the coefficient of t i.e. a_1_ in new cases data, is greater than the coefficient of t_2_, i.e. b_1_ than that of c_1_(a_1_>>b_1_>>c_1_) then with a smaller number of days we have to spend as lockdown to reduce the number. Again, if the coefficient of t^2^ i.e. b_1_ is less than c_1_ then we have to spend more days in lockdown to minimize the number. The same can also applicable for N_2_(t) and N_3_(t) also.

Suppose t = 10, then t^2^ =100 and t^3^ = 1000, we can stay in lockdown for 100 days but not the 1000 days. Hence the coefficient of t^3^ must be very less. Hence our approximation can be a valid and good approximation. Now to understand the minimization of growth to control the spreading of COVID-19 we have to find the minima of the dynamical equation (2). To do that let us first put the polynomial expansion of N_1_(t), N_2_(t) and N_3_(t) in the equation (1).

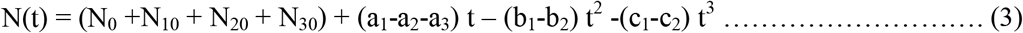

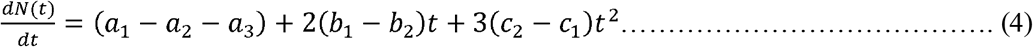

The equation (4) is actual dynamical equation of COVID 19 spreading in terms of new cases, recovery and death parameters. Zeroing the first order derivative of the equation (4) gives the no of days to reach extrema.

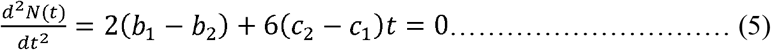

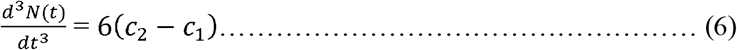

The equation (5) gives 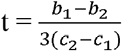. Now to equation (4) to be minimum (c_2_-c_1_) > 0, or **c_2_ > c_1_**.

As we have already discussed above that the coefficients c_i_ are related to long term case detection and recovery. This in equality suggest us that if the cubic power recovery coefficient is greater than the cubic power infected coefficient, then we can minimize the growth of spreading corona virus. This inherently implies that as the days go on the greater number of recoveries of the patient is required to overcome this problem. Simplest way it can be said that if we can increase the recovery rate gradually then we can easily reach the minima and will be able to control the COVID-19 epidemic. In case of India the recovery rate is gradually increasing compared to other countries. Hence it is possible control COVID 19 pandemic in India earlier than other countries.

### Data Analysis

To interpret our model, the daily affected case, recovery number and the mortality number with time are presented below. All the data are considered from 1 month of lock down to 49 days of lockdown. We only consider the data for India to illustrate our model. Fig 1 depicts the 3rd degree polynomial fit for daily new cases, recoveries and death cases of India. The coefficients of fitting are listed below in table 1. It is clear from the fitting that the coefficient of t^3^ in new case is less than the coefficient of t^3^ in recovery case. Hence the corona virus spreading can be controlled in India in due course of time.

**Fig 1:**
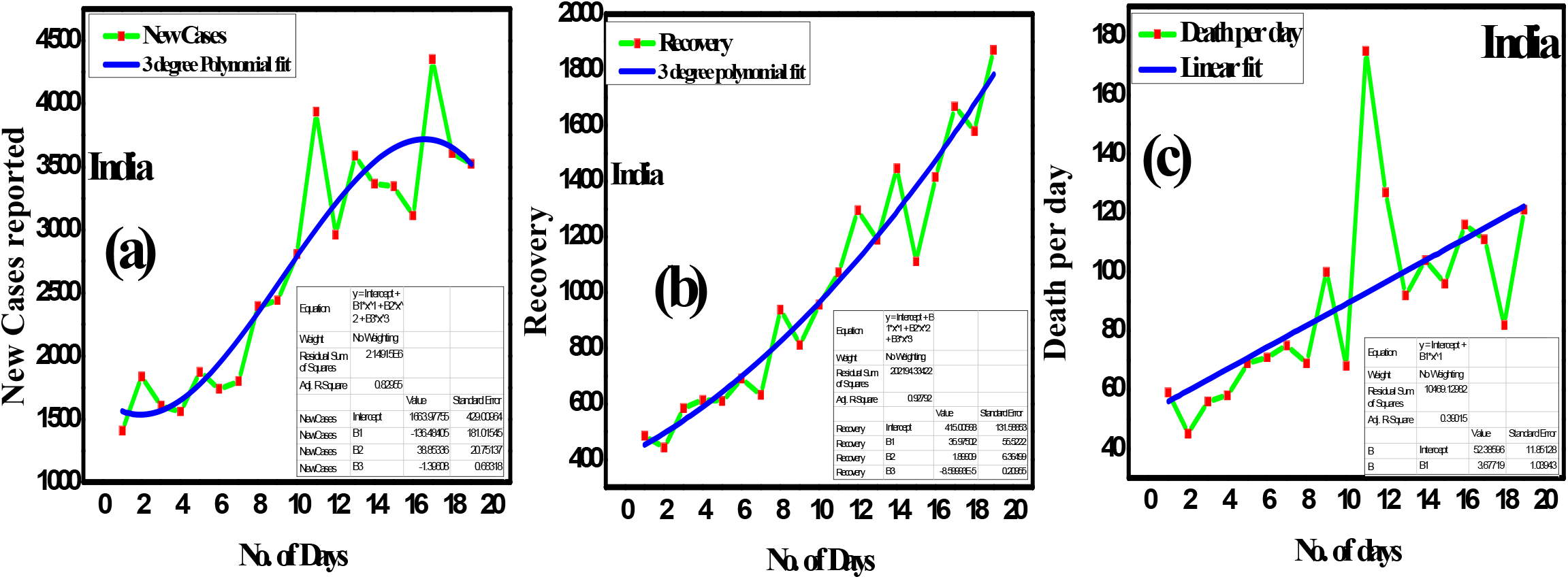
(a) new cases per day (b) recovery per day (c) death per day

**Table 1:** Coefficients of fitting for new cases, recovery and death in India

| India | Coefficient of $t$ | Coefficient of $t^2$ | Coefficient of $t^3$ |
| --- | --- | --- | --- |
| New Cases | -136.48405 | 38.85336 | -1.39608 |
| Recovery | 35.90572 | 1.89909 | $-8.59993 \times 10^{-5}$ |
| Death | 3.677 | NA | NA |

It is clear from the Fig 1 and Table 1 that to the coefficients of different term play an important role for controlling the spread of COVID 19. It is therefore clear that if we can maintain this condition and improve further then we can control COVID-19 in India. This can be achieved by a rigorous study of medicine that are used for treatment of corona virus.

### Relation of Recovery with Medicine

There is no such medication to CoVID-19 affected people. Firstly Hydroxychloroquine (HCQ) is seemed to be a good medication for such decease. India is a very popular source of such medicine. SARS-CoV-2 utilizes surface receptor like Angiotensin Converting Enzyme (ACE2) like CoVID-19 and hence it is believed that like chloroquine it can also interfere with ACE2 receptor glycosylation and thus prevents SARS-CoV2 attachment to the target cells. Moreover, HCQ is more potent agent than chloroquine due to less toxic nature. Chinese researchers found that chloroquine to be highly effective in reducing viral replication that can be easily achievable with standard dosing due to its favourable penetration in tissues including the lung [12]. Recently an excellent recovery was observed in 68% of the patients treated with one dose of remdisivir daily, according to an analysis published in The New England Journal of Medicine [13]. The recovery takes 18 days of time with single medication and the mortality rate is also very less in such case. Remdisivir is medication of Ebola virus where as HCQ is a for treatment of Rheumatoid Arthurites (RA). In a recent study in Gilead reveals that the average time to clinical improvement was 10 days in the five-day treatment group and 11 days in the 10-day treatment group [14]. More than half of the patients in both groups were discharged from the hospital by day 14. Hence Remdisivir is supposed to be considered as fast recovering medicine for the age group of 30 to 80 years. HCQ application to CoVID-19 patient takes a few days more to recover than Remdisivir ever after 5 days treatment [15]. Hence, Remdisivir and HCQ are both good medicine for CoVID-19 affected people. WHO has started a solidarity trail on 18^th^ March this year on CoVID-19 affected people using four medicine [16], 1. Remdisivir, 2. Lopanavir/ Ritonavir 3. Lopanavir and Ritonavir with interfirm beta and lastly 4. HCQ. The National Institute of Allergy and Infection Decease (NAID) has announced Remdisivir to be superior than HCQ. So, we suggest that Remdisivir can also be applicable for India for quicker recovery. Moreover, due to other vaccination (BCG, Hepatitis etc.) among younger age, the immunity power is very good and hence the recovery is faster (till now almost 40%). Wide coverage of BCG vaccination in India may lead to lesser fatal outcome of the decease but researchers are still working on it and relevant research papers are yet to be published. Since India is a tropical country, lots of people use Sunscreen Gel which are full of Zinc Oxide. The Zinc has excellent power to destroy the replication chain of CoVID 19. Hence it is also an important factor to stop spreading the Corona Virus.

### Conclusion

In this paper we have tried to establish a connection between the spreading of corona virus and recovery of corona virus. We use Indian statistical data to establish our theory. The effect of medication on different age of people is also discussed. The scope of further improvement of recovery rate in India is also suggested in terms of application of medicine to COVID 19 affected people. Hence if the recovery is faster, then the coefficient of t^3^ of recovery will be greater and hence we will have control over CoVID 19. We invite the researchers from all communities to come forward and work on COVID 19 according to their expertise. It may be helpful for fighting against such epidemic disease. Our analysis could give well explanation of daily affected people in last 21 days lockdown and in near future. We strongly believe that India will recover soon from the present situation.

## Data Availability

All the Data are taken from Worldometer website.

## Acknowledgement

JM thank to Dr. Dipak Bhowmik, IIT Kanpur for valuable reading of manuscript and Ms. Rinku Mondal, University of Burdwan for valuable suggestion.

## Authors Contribution

All the authors contribute equally throughout the data analysis, mathematical interpretation and paper writing.

